# Low levels of post-vaccination hemagglutination inhibition antibodies and their correlation with influenza protection among healthcare workers during the 2024/2025 A/H1N1 outbreak in Japan

**DOI:** 10.1101/2025.02.06.25321776

**Authors:** Shohei Yamamoto, Tetsuya Mizoue, Mugen Ujiie, Kumi Horii, Junko S. Takeuchi, Maki Konishi, Wataru Sugiura, Norio Ohmagari

## Abstract

**Background:** After the prolonged COVID-19 pandemic, during which the seasonal influenza epidemic was suppressed, Japan experienced a record-breaking influenza A/H1N1 outbreak in the 2024/2025 season. This situation also raises a concern about the immunogenicity of the annual quadrivalent inactivated influenza vaccine (QIIV). This study evaluated post-vaccination hemagglutination inhibition (HI) antibody titers and their association with influenza infection risk among healthcare workers.

**Methods:** A serosurvey was conducted among staff at a national medical and research center in Tokyo in December 2024, one month after staff received the QIIV. HI antibody titers against vaccine strains were measured, and participants were followed for influenza infection until January 2025. Seroprotection was defined as an HI titer ≥40. A Cox proportional hazards model assessed the association between HI titers and infection risk among vaccinated participants.

**Results:** Among 1,507 vaccinated participants, only 12.7% had seroprotective HI titers against A/H1N1. Around 90% had no influenza history for at least four seasons and had received repeated vaccinations over two seasons. Participants with HI titers <40 had a 4-fold higher infection risk than those with titers ≥40. A dose-response association was observed, even within the range below the titer of 40. Relative to titers <10, titers of 10 and 20 conferred 47.3% and 57.9% protection, respectively.

**Conclusions:** After a prolonged period without a major influenza epidemic, HI titers against A/H1N1 were extremely low in vaccinated healthcare workers. Nonetheless, higher post-vaccination HI titers, even at relatively low levels, were associated with protection, supporting the benefit of vaccines.

**Main points:** During Japan’s record-breaking 2024/2025 influenza A/H1N1 outbreak, HI antibody titers against A/H1N1 were extremely low in vaccinated healthcare workers. Nonetheless, even relatively low post-vaccination HI titers conferred moderate protection, highlighting the benefits of vaccination despite reduced immunogenicity.

## Introduction

During the 2024/2025 season, Japan recorded the highest number of influenza cases per week since 1999, when the country began tracking the number [1]. The majority of the reported cases were due to the A/H1N1 sublineage strain. This ongoing large-scale outbreak is likely due to the lack of herd immunity to influenza after the long-term COVID-19 pandemic. Preventive measures against COVID-19, such as mask-wearing and social distancing, have been highly effective in reducing influenza virus transmission, substantially suppressing the influenza epidemic in Japan from the 2020/2021 to 2021/2022 seasons [1]. Tokyo’s annual serosurveys have shown a substantial decrease in the prevalence of seroprotection against the A/H1N1 strain, defined as hemagglutination inhibition (HI) antibody titers ≥40, among unvaccinated individuals, from 98.4% in 2019 to 2.7% in 2023 [2].

In a situation where herd immunity against influenza is declining, vaccine-induced immunity is critical. Before the COVID-19 pandemic, clinical trials in adults assessed the immunogenicity of the inactivated quadrivalent influenza vaccines (QIIV), reporting an increase in the prevalence of seroprotection from 20–70% pre-vaccination to 80–100% post-vaccination [3]. As these results indicate, annual vaccination programs are designed to boost pre-existing immunity that was already primed before vaccination [4]. Given the current situation in Japan where pre-vaccination immunity is extremely low, concerns arise about whether vaccination can induce immunity as effectively as before the COVID-19 pandemic. So far, data on vaccine immunogenicity following the long-term COVID-19 pandemic remain limited: one available report from Tokyo’s annual serosurvey in the last 2023/2024 season reported 4.0% of 101 vaccinated individuals had an HI titer ≥40 against A/H1N1 [2]. In addition, it remains elusive whether such vaccine-induced antibody titers correlate with protection against influenza infection during the 2024/2025 A/H1N1 outbreak.

In the National Center for Global Health and Medicine (NCGM) in Tokyo, the staff received the QIIV as an employee immunization program in October and November 2024; one month later, approximately 1600 staff participated in a serosurvey assessing influenza HA antibodies; shortly after, a major influenza outbreak occurred. This situation prompted us to evaluate HI antibody levels and their related factors in vaccinated healthcare workers and examine the association between pre-infection HI antibody titers and the risk of influenza infection early in the 2024/2025 season.

## Methods

### Study setting and population

The NCGM is a national medical and research center for specific areas, including infectious diseases. As an annual influenza vaccination program, the staff received the QIIV vaccine (DENKA Co., Ltd.) against A/Victoria/4897/2022 (H1N1), A/California/122/2022 (H3N2), B/Austria/1359417/2021 (Victoria lineage), and B/Phuket/3073/2013 (Yamagata lineage) in October and November 2024. This vaccine, along with all influenza vaccines approved for adults in Japan, is a non-adjuvanted inactivated vaccine containing the four antigens.

In the NCGM, a repeated serological study was launched in July 2022 to monitor the spread of SARS-CoV-2 infection among staff during the COVID-19 epidemic. The details of this study have been reported elsewhere [5, 6]. With the circulation of seasonal influenza returning to pre-COVID-19 pandemic levels, we added monitoring the spread of influenza infections in 2023. In the eleventh serosurvey on December 2–6, 2024, we measured HI antibody titers to influenza and asked participants for a history of influenza diagnosis and vaccination via a questionnaire, which was confirmed by records of the infection control unit and the labor-management department, respectively. Written informed consent was obtained from all participants. This study was approved by the NCGM Ethics Committee (approved number: NCGM-G-003598). Of 1918 staff invited to the eleventh serosurvey, 1609 (84%) participated. Of these, we excluded those without data on vaccination for the 2024/2025 season (n=1) and those diagnosed with influenza during the current season before the serosurvey (n=10), leaving 1598 participants for analyses.

### Antibody testing

The serum HI antibody titers against the four influenza virus antigens, which correspond to the antigens of the 2024/2025 seasonal QIIV in Japan, were measured at an external laboratory (LSI Medience Corporation, Tokyo) following the standard World Health Organization reagent preparation protocols [7]. These antigens for the assays were sourced from DENKA Co., Ltd., the same manufacturer of the 2024/2025 seasonal vaccine administered at NCGM. The assay used goose red blood cells (RBCs) for A/H3N2 and chicken RBCs for the other antigens. Serum-virus incubation was conducted at 37°C to ensure optimal sensitivity and specificity for the vaccine antigens. Serum samples were tested in serial 2-fold dilutions with a starting dilution of 1:10. The HI antibody titer was defined as the reciprocal of the highest serum dilution that gave a positive result. The lower limit of quantitation was a titer of 10, and samples with titers below this threshold were assigned a titer of 5.

### Ascertainment of subsequent Influenza infection

To assess the association between post-vaccination HI titers against A/H1N1 and the risk of breakthrough infection, we followed the participants for influenza incidence from the date of the survey participation (December 2–6, 2024) through January 15, 2025, during which A/H1N1 was predominant in Japan. We identified patients with influenza using in-house registry data that included information on the date of diagnosis, virus type (A or B), symptoms, and hospitalization. As per the NCGM rule, staff should undergo diagnostic tests for influenza and COVID-19 when they have cold-like symptoms, and if the test results are positive, they must report the results to the NCGM Hospital Infection Prevention and Control Unit.

### Data on the number of influenza cases and the distribution of influenza strains and clades across Japan

We obtained data on the number of influenza cases per sentinel in Japan for the 2019/2020 to 2024/2025 seasons from the Infectious Disease Weekly Report (IDWR) [1] published by the National Institute of Infectious Diseases (NIID), Japan. We also collected data on influenza virus isolations and detections from the 2019/2020 to 2024/2025 seasons in Japan from the Infectious Agents Surveillance Report (IASR) [8] published by the NIID. The A/H1N1 clade distribution from the 2019/2020 to the 2024/2025 seasons in Japan was analyzed using domestic data from the Global Initiative on Sharing All Influenza Data (GISAID) EpiFlu database (https://gisaid.org/) as of January 27, 2025.

### Statistical Analysis

We defined seroprotection as HI titer ≥40, a threshold generally accepted to correspond to a 50% reduction in the risk of contracting influenza [9]. Using a robust Poisson regression, we calculated the prevalence of seroprotection with 95% confidence intervals (CIs) based on the 2024/2025 vaccination status. We compared geometric mean titers (GMT) of HI antibodies according to the 2024/2025 vaccination status using linear regression, with HI titers analyzed on a log scale. Among the participants vaccinated for the 2024/2025 season, we compared the GMTs by the history of influenza diagnosis in the 2022/2023 or 2023/2024 seasons (no diagnosis, diagnosed with A, or diagnosed with B) and by prior vaccination status (2024/2025 season only vs. both 2023/2024 and 2024/2025 seasons) with adjustment for age and sex. In the analysis of prior vaccination status, we excluded those with a history of influenza diagnosis in the 2022/2023 or 2023/2024 seasons.

We calculated the person-time from the date of the serosurvey participation through the date of influenza diagnosis, additional vaccination, resignation (retirement or leave of absence), or January 15, 2025, whichever occurred first. Among the participants vaccinated for the 2024/2025 season, we fitted a Cox proportional hazards regression model to examine the association between HI titer against A/H1N1 and the risk of breakthrough infection while adjusting for age (continuous) and sex (male, female). The estimated hazard ratio (HR) was used to calculate the protection (%) according to the formula: (1 – HR) × 100. The proportional-hazards assumption was evaluated using Schoenfeld residuals, and no violation of the assumption was found for any covariate (p > 0.05 for all). Statistical analyses were performed with Stata version 18.0 (StataCorp), and graphics were generated with Prism version 9 (GraphPad). All P values were 2-sided, and statistical significance was set at P < 0.05.

## Results

### Baseline characteristics

Of the 1598 participants, 72.1% were female, and the median age [interquartile range: IQR] was 38 [27–50] years (**Table 1**). The most common occupations were nurses (37.5%), followed by allied healthcare workers (15.4%), researchers (15.1%), administrative staff (14.6%), and doctors (13.1%). The proportion of QIIV vaccine recipients was 94.3% and 87.7% for the current 2024/2025 season and the previous 2023/2024 season, respectively. The median interval [IQR] between the 2024/2025 vaccination to the serosurvey was 29 [26–38] days. A total of 123 participants (7.7%) had a history of influenza diagnosis in the 2022/2023 season (n=25) and/or the 2023/2024 season (n=99).

**Table 1.** Participants characteristics.

|  | **Total** |
| --- | --- |
|  | N=1598 |
| **Sex** |  |
| Male | 446 (27.9) |
| Female | 1152 (72.1) |
| **Age** | 38 [27–50] |
| 20–29 | 527 (33.0) |
| 30–39 | 323 (20.2) |
| 40–49 | 323 (20.2) |
| ≥50 | 425 (26.6) |
| **Job** |  |
| Doctors | 209 (13.1) |
| Nurses | 600 (37.5) |
| Allied healthcare professionals | 246 (15.4) |
| Administrative staff | 233 (14.6) |
| Researcher | 242 (15.1) |
| Others | 68 ( 4.3) |
| **History of influenza vaccination** |  |
| 2023/24 season | 1,376 (87.7) |
| 2024/25 season | 1,507 (94.3) |
| Days since the 2024/25 season vaccination | 29 [26–38] |
| **History of influenza diagnosis** |  |
| 2022/23 season | 25 (1.6) |
| 2023/24 season | 99 (6.1) |
| 2022/23 or 2023/24 seasons | 123 (7.7) |

Data are presented as numbers (%) for categorical variables and median [interquartile range] for continuous variables.
Missing values: 2023/24 season vaccination status (n=29)

### Prevalence of seroprotection and HI titers across the 2024/2025 vaccination status

Figure 1. shows the comparison of the HI antibodies by the 2024/2025 season vaccination status. The prevalence of seroprotection was generally low but higher in the vaccinated group compared to the unvaccinated group for all four strains: A/H1N1 (12.7% vs. 5.5%), A/H3N2 (68.1% vs. 25.3%), B/Victoria (29.3% vs. 9.9%), and B/Yamagata (40.9% vs. 25.3%). The ratios of GMT(95% CIs) of HI antibodies for the vaccinated to unvaccinated group were 1.4 (1.2–1.7) for A/H1N1, 3.0 (2.5–3.7) for A/H3N2, 2.0 (1.7–2.4) for B/Victoria, and 1.7 (1.4–2.0) for B/Yamagata (all P < 0.001).

**Figure 1.**
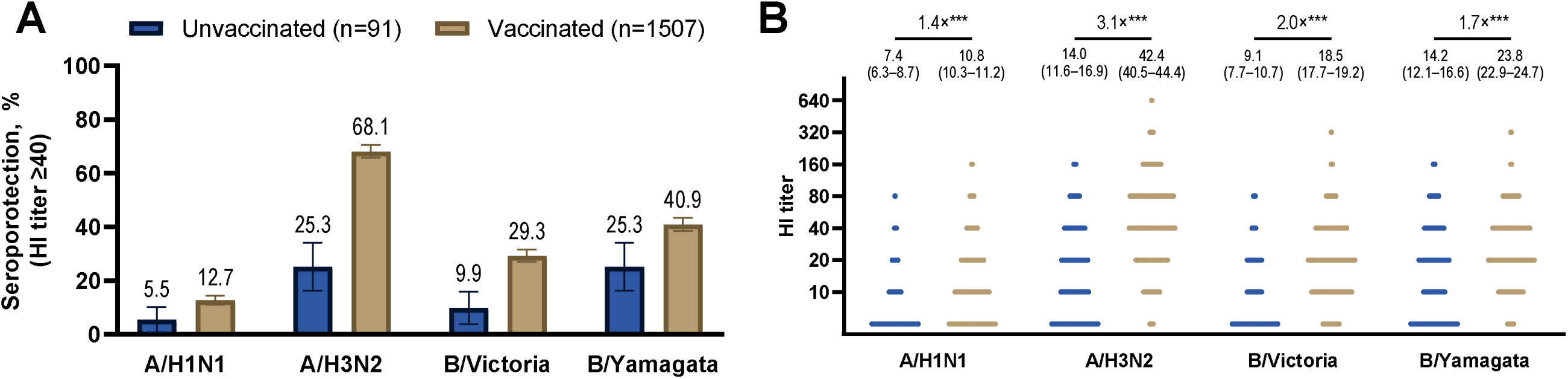
Prevalence of seroprotection (**A**) and comparison of the geometric mean hemagglutination inhibition titers (**B**) against four influenza virus strains by the 2024/2025 vaccination status. (**A**): The left panel displays the prevalence of seroprotection (HI titers ≥ 40), stratified by 2024/25 vaccination status. Bars represent the prevalence of seroprotection, with error bars indicating 95% CIs estimated using robust Poisson regression. (**B**): The right panel compares geometric mean HI titers by 2024/25 vaccination status. The presented values are geometric mean titers and 95% CIs, and the fold-change values (×) indicate the ratio of geometric mean titers, estimated using linear regression (***P < 0.001). Abbreviations: HI, hemagglutination inhibition; 95% CI, 95% confidence interval.

### HI titers across a history of influenza diagnosis

**Figure 2A** illustrates the comparison of HI antibody titers across the history of influenza diagnosis in the 2022/2023 or 2023/2024 seasons among participants who received the 2024/2025 season vaccine. Participants with a history of influenza A showed slightly higher GMTs against A/H1N1 compared to those without the history (12.1 vs. 10.7), yielding a GMT ratio of 1.13 (0.95–1.34). Their GMTs against A/H3N2 were significantly higher than those without the history (54.3 vs. 41.8), with a GMT ratio of 1.30 (1.10–1.53). Those with a history of influenza B had slightly higher GMTs against B/Victoria than those without the history (22.9 vs. 18.5), corresponding to a GMT ratio of 1.24 (0.84–1.81).

**Figure 2.**
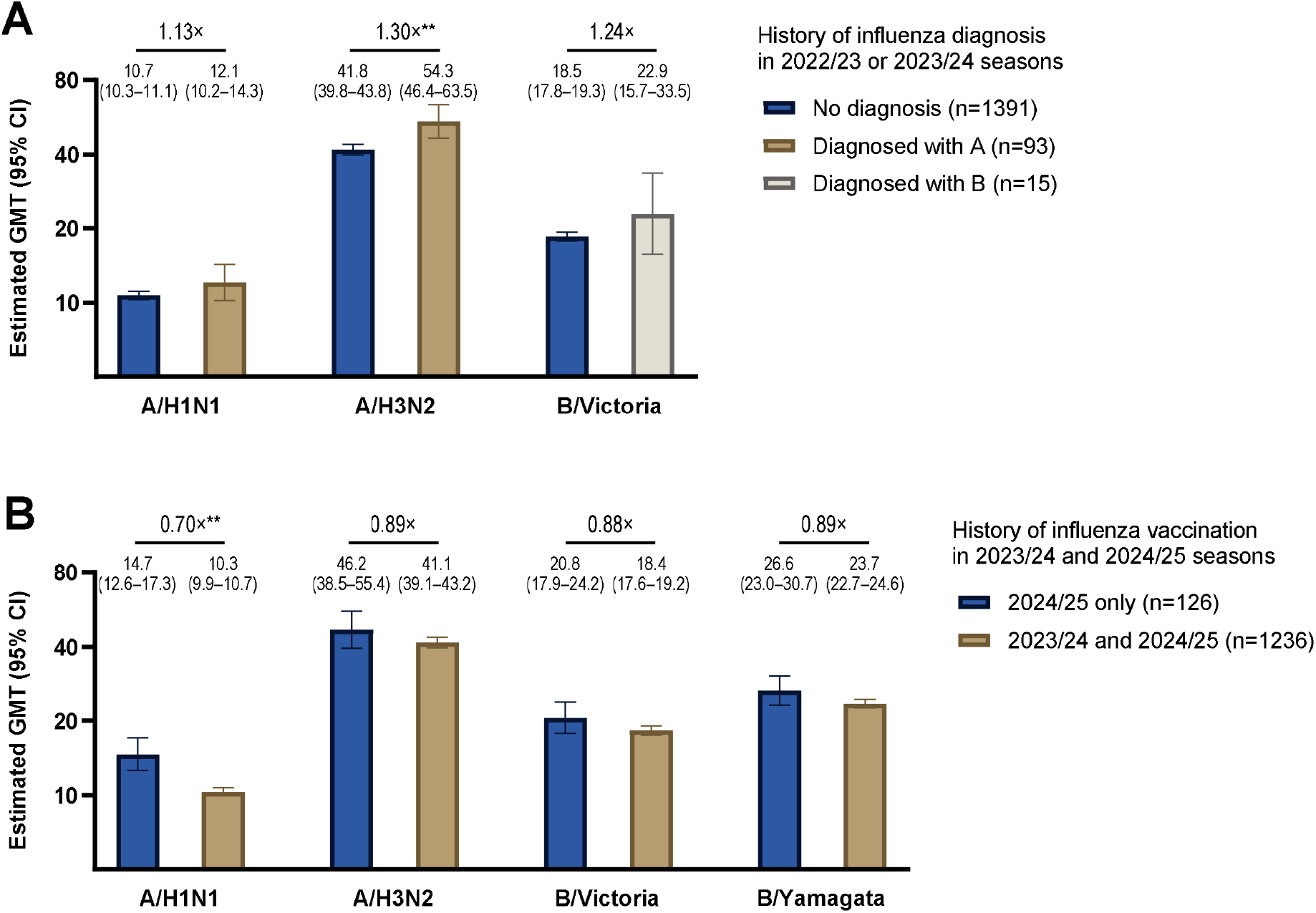
Comparison of the geometric mean hemagglutination inhibition titers by the history of influenza diagnosis in the 2022/2023 or 2023/2024 (A) and by the current and previous vaccination status (B) among the 2024/2025 vaccinated participants. (**A**): The upper panel compares the GMT by the history of influenza diagnosis in 2022/2023 or 2023/2024 among the 2024/2025 vaccinated participants. For HI titers against A/H1N1 and A/H3N2, we compared them between those without a history of influenza and those with a history of influenza A. For HI titers against B/Victoria, we compared the titer between those without a history of influenza and those with a history of influenza B. Since the B/Yamagata has not been detected since 2020, we did not compare that titer. Those who lacked information about the type of influenza (A or B) were excluded from this analysis (n=8). (**B**): The lower panel compares GMT by the 2024/2025 and 2023/2024 vaccination status among the 2024/2025 vaccinated participants. Those with a history of influenza diagnosis in 2022/2023 or 2023/2024 were excluded from this analysis (n=116). The bars indicate the estimated GMT, and the I-bars indicate 95% CIs. The presented values are GMT and 95% CIs, and the fold-change values (×) indicate the ratio of GMT, estimated using linear regression with adjustment for age and sex (*P < 0.05, **P <0.01, ***P < 0.001). Abbreviations: GMT, geometric mean titer; HI, hemagglutination inhibition; 95% CI, 95% confidence interval.

### HI titers across current and previous vaccination status

**Figure 2B** compares HI antibody titers based on influenza vaccination history in the current (2024/2025) and previous (2023/2024) seasons. Participants vaccinated in both seasons had lower GMTs for all four strains than those vaccinated only in the current season, with the greatest difference seen for A/H1N1. The GMT ratios were 0.70 (0.59–0.82) for A/H1N1, 0.89 (0.74–1.08) for A/H3N2, 0.88 (0.75–1.04) for B/Victoria, and 0.89 (0.77–1.03) for B/Yamagata.

### Influenza epidemics and detected strains across Japan in the early 2024/2025 season

In the 2024/2025 season in Japan, the nationwide epidemic began in week 44 (October 30–November 5, 2024) when cases per sentinel exceeded 1.00 (**Supplemental Figure 1**). During the period corresponding to the study follow-up (December 2, 2024–January 15, 2025), cases surged, reaching 9.03 in week 49 (December 4–10) and peaking at 64.39 in week 52 (December 25–31), the highest since the IDWR system began in 1999. Subsequently, cases declined to 33.82 in week 1 of 2025 (January 1–7), followed by 18.38 in week 3 (January 15–21). From week 49 of 2024 to week 3 of 2025, 634 samples were analyzed for their strains in Japan, and 599 (94.5%) were detected as A/H1N1(pdm09), followed by 27 (4.3%) as A/H3N2 and 8 (1.3%) as B/Victoria (**Supplemental Figure 2**). All A/H1N1 viruses belonged to clade 6B.1A.5a.2a based on the GISAID EpiFlu database (**Supplemental Figure 3**).

### Incidence of Influenza among vaccinated participants during the follow-up

During the A/H1N1 predominant follow-up period, 56 participants were diagnosed with influenza among vaccinated participants. The incidence rate was 9.0 per 10000 person-days. Of the 56 diagnosed individuals, 46 had identified the type of influenza virus: 45 (98%) were Influenza A, and 1 (2%) was Influenza B. All cases reported some symptoms at diagnosis: 53 (95%) had a fever of ≥ 37.5D, and 24 (43%) had upper respiratory symptoms. No cases required hospitalization.

### Association between HI titers against A/H1N1 and protection among vaccinated participants

Among vaccinated participants, the breakthrough infection rate was four times higher in those with pre-infection HI titers <40 compared to those with titers ≥40 (10.0 vs. 2.5 per 10,000 person-days) (**Table 2**). No cases were observed in the group with titers ≥80. The Cox regression analysis showed that protection against influenza infection increased steadily with increasing HI titer against A/H1N1. Compared to HI titer <10, the protection (95% CIs) of HI titer 10, 20, 40, ≥80 were 47.3% (1.2–71.8), 57.9% (7.5–80.9), 80.3% (18.2–95.3), and 100% (not applicable), respectively (P for trend < 0.01).

**Table 2.**
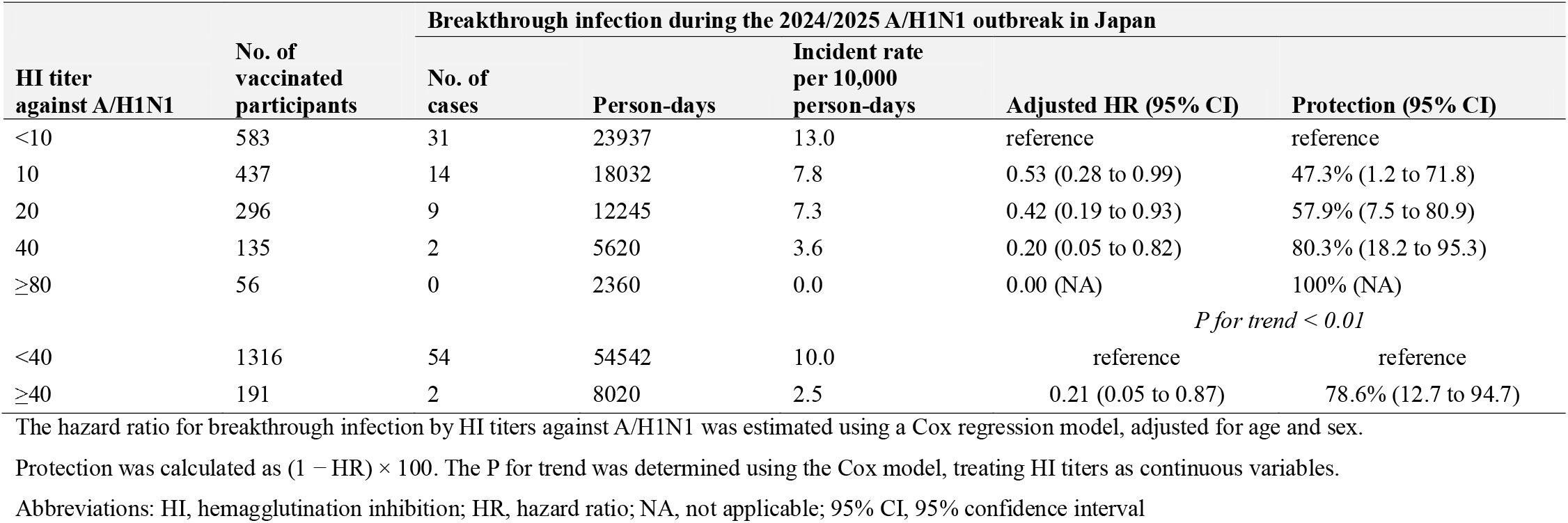
Association between pre-infection HI titers against A/H1N1 and the protection against influenza infection among vaccinated participants during the 2024/25 A/H1N1 outbreak in Japan.

## Discussion

In December 2024, the prevalence of seroprotection was low, especially for A/H1N1, even among healthcare workers who received the vaccine. Nevertheless, there was a dose-response relationship between post-vaccination HI titers against A/H1N1 and influenza protection during the high circulation period of A/H1N1, showing significant protection even at HI titers of 10 and 20.

Our finding of 12.7% of seroprotection against A/H1N1 among vaccinated individuals in the 2024/2025 season was substantially lower than the results of the 2009–2011 clinical trials on the immunogenicity of QIIV (80–100%) [3], while it is compatible with a low prevalence among vaccinated Tokyo residents in the 2023/2024 season (4.0%) [2]. These results suggest that the immunogenicity of the influenza vaccine has decreased during the long-term COVID-19 pandemic. This may be due, at least in part, to a decline in influenza-specific immune memory over the extended period without a major influenza epidemic. Current inactivated influenza vaccines without adjuvants cannot induce robust T cell-dependent memory B cells (MBCs) [10], which play a crucial role in antibody production [11]. Thus, the antibody response to these vaccines primarily depends on pre-existing MBCs [12, 13]. Natural influenza infection can strongly boost MBCs, although this memory fades with time [14]. Most of the present study participants (92.3%) were not diagnosed with influenza during the last four seasons (2020/2021 to 2023/2024). In addition, 6% of the vaccinated participants had contracted influenza A within the last two seasons, and they had significantly higher HI titers against A/H3N2 than those without a recent infection history but only marginally higher against A/H1N1, indicating that most previous influenza A cases were likely due to A/H3N2. These findings suggest that nearly all participants may not have had the opportunity to enhance their influenza A/H1N1-specific immune memory for several seasons.

Another possible reason for the low immunogenicity is repeated annual vaccination. In this study, approximately 90% of participants received influenza vaccine in both the current and previous seasons, and they had 11-30% lower HI titers than those vaccinated only in the current season. This finding is consistent with previous immunologic studies [15, 16] and is compatible with vaccine effectiveness (VE) studies showing a 7-18% lower VE against influenza infection in individuals vaccinated in both seasons compared to those vaccinated only in the current season [17]. This phenomenon could be explained by the antigenic distance hypothesis [18]; pronounced negative interference occurs when previous and current vaccine strains are antigenically similar. The 2023/2024 and 2024/2025 QIIV vaccines used for the NCGM in-house vaccination program contained the same antigens except for A/H3N2 (**Supplementary Table 1**). Notably, A/H3N2—the only strain that changed from the previous season—elicited the highest antibody titer after vaccination compared to the other three antigens (GMTs: 42.4 vs. 10.8-23.8). However, these data should not be interpreted as evidence against vaccination can be skipped. According to VE studies [19-21], vaccination in both the current and previous seasons showed higher effectiveness against influenza-related hospitalizations than vaccination in either season alone, supporting current recommendations for annual influenza vaccination.

Among vaccinated participants, we found that a higher HI titer against A/H1N1 was associated with greater protection against infection during the A/H1N1 outbreak in a dose-response manner. This finding is reasonable because the A/H1N1 antigen of the 2024/2025 vaccine was antigenically well-matched to the current circulating virus in Japan (clade: 6B.1A.5a.2a) [22]. An HI titer of 40 has been accepted as a threshold corresponding to a 50% reduction in the risk of contracting influenza [9]. However, we found that an HI titer of 10–20 against A/H1N1 conferred 47.3%–57.9% protection against infection, which is consistent with a meta-analysis showing that the HI titer corresponding to 50% protection is as low as 17 (95% CI: 9–28) [23]. In the present study, 61.3% of vaccinated individuals had HI titers ≥10 (**Supplemental Figure 4**); thus, more than half of vaccine recipients may have achieved moderate protection.

This study boasts several key strengths, including a relatively large cohort, a high participation rate of 84%, and the availability of HI antibody titers measured around 4 weeks after the vaccination and immediately prior to the influenza outbreak. We should also acknowledge limitations. First, we did not measure pre- and post-vaccination HI titers from the same participants, preventing within-subject comparison and limiting the accurate immune response to vaccination assessment. Second, we did not collect a history of influenza diagnosis before the COVID-19 pandemic. Although pre-COVID-19 studies suggested that natural infection-induced antibody immunity to HA is long-lived (over 50 years) [24], our data did not clarify whether this remains the case following a prolonged actual absence of influenza exposure, including asymptomatic infections. Third, while information on the type of influenza (A or B) was available in both the history and follow-up surveys, data on specific subtypes, such as A/H1N1 and A/H3N2, were not collected in this study. Regarding history, since both A/H1N1 and A/H3N2 predominated in the past two seasons (**Supplemental Figure 2**), it is not possible to estimate which subtype virus represents the history of influenza A. For the follow-up survey, the cases were most likely due to A/H1N1, which accounted for 94.5% of sequenced influenza samples in Japan during the follow-up (**Supplemental Figure 2**). Fourth, we did not assess other humoral or cell-mediated immune indicators. For example, neutralizing antibodies directly reflect the functional ability to block viral infection [24], and cell-mediated immune responses are crucial for long-term and cross-protective immunity [11]. Fifth, we did not conduct active surveillance of influenza infection during the follow-up, possibly underestimating the total cases. Finally, this study included only healthcare workers from a single medical center in Tokyo, most of whom received annual influenza vaccinations and had experienced fewer infections over at least four seasons. In addition, no severe cases were observed. Caution should be exercised when generalizing our findings to different populations, particularly those at high risk of severe influenza, such as children, older adults, and patients with immunosuppressive conditions.

In conclusion, HI titers against A/H1N1 were extremely low even in vaccinated healthcare workers after a prolonged period without a major influenza epidemic. Nevertheless, higher post-vaccination HI titers, even at modest levels below 40, were associated with greater protection against influenza infection, supporting the benefit of influenza vaccination in the post-COVID-19 era. Future studies should explore the mechanisms behind the low immunogenicity after vaccination and address improving the immunological response and effectiveness of the influenza vaccine.

## Data Availability

All data produced in the present study are available upon reasonable request to the authors.

## Author Contributions

Drs. Yamamoto and Mizoue had full access to all data in the study and took responsibility for the integrity of the data and the accuracy of the data analysis.

Conceptualization: SY and TM. Methodology: SY and TM. Software: SY, JT, and MK. Validation: MK and TM. Formal analysis: SY. Investigation: SY, TM, KH, JT, and MK. Resources: TM, KH, JT, WS, and NO. Data Curation: SY, JT, and MK. Writing - Original Draft: SY and TM. Visualization: SY and JT. Supervision: TM, MU, and NO. Project administration: SY and TM. Funding acquisition: SY and TM. All authors contributed to manuscript revisions and editing.

## Additional Contributions

We thank Mika Shichishima for her contribution to data collection.

## References

1. National Institute of Infection Diseases. Infectious Disease Weekly Report JAPAN (IDWR). Available at: https://www.niid.go.jp/niid/en/idwr-e.html. Accessed January 15 2025.

2. Tokyo Metropolitan Infectious Disease Surveillance Center. Epidemiological Surveillance of Vaccine-Preventable Diseases. Available at: https://idsc.tmiph.metro.tokyo.lg.jp/vpd/. Accessed January 15 2025.

3. Moa AM, Chughtai AA, Muscatello DJ, Turner RM, MacIntyre CR. Immunogenicity and safety of inactivated quadrivalent influenza vaccine in adults: A systematic review and meta-analysis of randomised controlled trials. Vaccine 2016; 34:4092–102.

4. Palgen J-L, Feraoun Y, Dzangué-Tchoupou G, et al. Optimize Prime/Boost Vaccine Strategies: Trained Immunity as a New Player in the Game. Frontiers in Immunology 2021; 12.

5. Yamamoto S, Oshiro Y, Inamura N, et al. Durability and determinants of anti-SARS-CoV-2 spike antibodies following the second and third doses of mRNA COVID-19 vaccine. Clin Microbiol Infect 2023.

6. Yamamoto S, Matsuda K, Maeda K, et al. Preinfection Neutralizing Antibodies, Omicron BA.5 Breakthrough Infection, and Long COVID: A Propensity Score-Matched Analysis. The Journal of Infectious Diseases 2023.

7. Organization WH. WHO manual on animal influenza diagnosis and surveillance: World Health Organization, 2002.

8. National Institute of Infectious Diseases. Infectious Agents Surveillance Report (IASR). Available at: https://www.niid.go.jp/niid/ja/iasr-inf.html. Accessed January 27 2025.

9. Hobson D, Curry RL, Beare AS, Ward-Gardner A. The role of serum haemagglutination-inhibiting antibody in protection against challenge infection with influenza A2 and B viruses. Epidemiology and Infection 1972; 70:767–77.

10. Abreu R, Ross TM. Influenza – A new pathogen every year. Current Opinion in Systems Biology 2018; 12:12–21.

11. Syeda MZ, Hong T, Huang C, Huang W, Mu Q. B cell memory: from generation to reactivation: a multipronged defense wall against pathogens. Cell Death Discovery 2024; 10.

12. Cheng X, Zengel JR, Suguitan AL, et al. Evaluation of the Humoral and Cellular Immune Responses Elicited by the Live Attenuated and Inactivated Influenza Vaccines and Their Roles in Heterologous Protection in Ferrets. The Journal of Infectious Diseases 2013; 208:594–602.

13. Abreu RB, Kirchenbaum GA, Clutter EF, Sautto GA, Ross TM. Preexisting subtype immunodominance shapes memory B cell recall response to influenza vaccination. JCI Insight 2020; 5.

14. Kurosaki T, Kometani K, Ise W. Memory B cells. Nature Reviews Immunology 2015; 15:149–59.

15. Thompson MG, Naleway A, Fry AM, et al. Effects of Repeated Annual Inactivated Influenza Vaccination among Healthcare Personnel on Serum Hemagglutinin Inhibition Antibody Response to A/Perth/16/2009 (H3N2)-like virus during 2010-11. Vaccine 2016; 34:981–8.

16. Ng TWY, Perera RAPM, Fang VJ, et al. The Effect of Influenza Vaccination History on Changes in Hemagglutination Inhibition Titers After Receipt of the 2015–2016 Influenza Vaccine in Older Adults in Hong Kong. The Journal of Infectious Diseases 2020; 221:33–41.

17. Jones-Gray E, Robinson EJ, Kucharski AJ, Fox A, Sullivan SG. Does repeated influenza vaccination attenuate effectiveness? A systematic review and meta-analysis. The Lancet Respiratory Medicine 2023; 11:27–44.

18. Smith DJ, Forrest S, Ackley DH, Perelson AS. Variable efficacy of repeated annual influenza vaccination. Proceedings of the National Academy of Sciences 1999; 96:14001–6.

19. Cheng AC, Macartney KK, Waterer GW, et al. Repeated Vaccination Does Not Appear to Impact Upon Influenza Vaccine Effectiveness Against Hospitalization With Confirmed Influenza. Clinical Infectious Diseases 2017; 64:1564–72.

20. Rondy M, Launay O, Castilla J, et al. Repeated seasonal influenza vaccination among elderly in Europe: Effects on laboratory confirmed hospitalised influenza. Vaccine 2017; 35:4298–306.

21. Hsu P-S, Lian I-B, Chao D-Y. A Population-Based Propensity Score-Matched Study to Assess the Impact of Repeated Vaccination on Vaccine Effectiveness for Influenza-Associated Hospitalization Among the Elderly. Clinical Interventions in Aging 2020; Volume 15:301–12.

22. National Institute of Infectious Diseases. Influenza virus epidemic strains Antigenic analysis and genetic phylogenetic tree in the 2024/2025 season, 2024.

23. Coudeville L, Bailleux F, Riche B, Megas F, Andre P, Ecochard R. Relationship between haemagglutination-inhibiting antibody titres and clinical protection against influenza: development and application of a bayesian random-effects model. BMC Medical Research Methodology 2010; 10:18.

24. Krammer F. The human antibody response to influenza A virus infection and vaccination. Nature Reviews Immunology 2019; 19:383–97.

