## Supplemental Materials for "Low levels of post-vaccination hemagglutination inhibition antibodies and their correlation with influenza protection among healthcare workers during the 2024/2025 A/H1N1 outbreak in Japan"

**Supplemental Table 1**. The influenza vaccine strains used in Japan from the 2018/2019 to the 2024/2025 seasons

| Season | A(H1N1)pdm09-like Virus | A(H3N2)-like Virus | B/Victoria Lineage-like Virus | B/Yamagata Lineage-like Virus |
|---|---|---|---|---|
| 2019/2020 | A/Brisbane/02/2018 (IVR-190) | A/Kansas/14/2017 (X-327) | B/Maryland/15/2016 (NYMC BX-69A) | B/Phuket/3073/2013 |
| 2020/2021 | A/Guangdong-Maonan/SWL1536/2019 (CNIC-1909) | A/Hong Kong/2671/2019 (NIB-121) | B/Victoria/705/2018 (BVR-11) | B/Phuket/3073/2013 |
| 2021/2022 | A/Victoria/1/2020 (IVR-217) | A/Tasmania/503/2020 (IVR-221) | B/Victoria/705/2018 (BVR-11) | B/Phuket/3073/2013 |
| 2022/2023 | A/Victoria/1/2020 (IVR-217) | A/Darwin/9/2021 (SAN-010) | B/Austria/1359417/2021 (BVR-26) | B/Phuket/3073/2013 |
| 2023/2024 | A/Victoria/4897/2022 (IVR-238) | A/Darwin/9/2021 (SAN-010) | B/Austria/1359417/2021 (BVR-26) | B/Phuket/3073/2013 |
| 2024/2025 | A/Victoria/4897/2022 (IVR-238) | A/California/122/2022 (SAN-022) | B/Austria/1359417/2021 (BVR-26) | B/Phuket/3073/2013 |

The red font indicates that the strain is the same as the vaccine production strain for the 2024/2025 season.

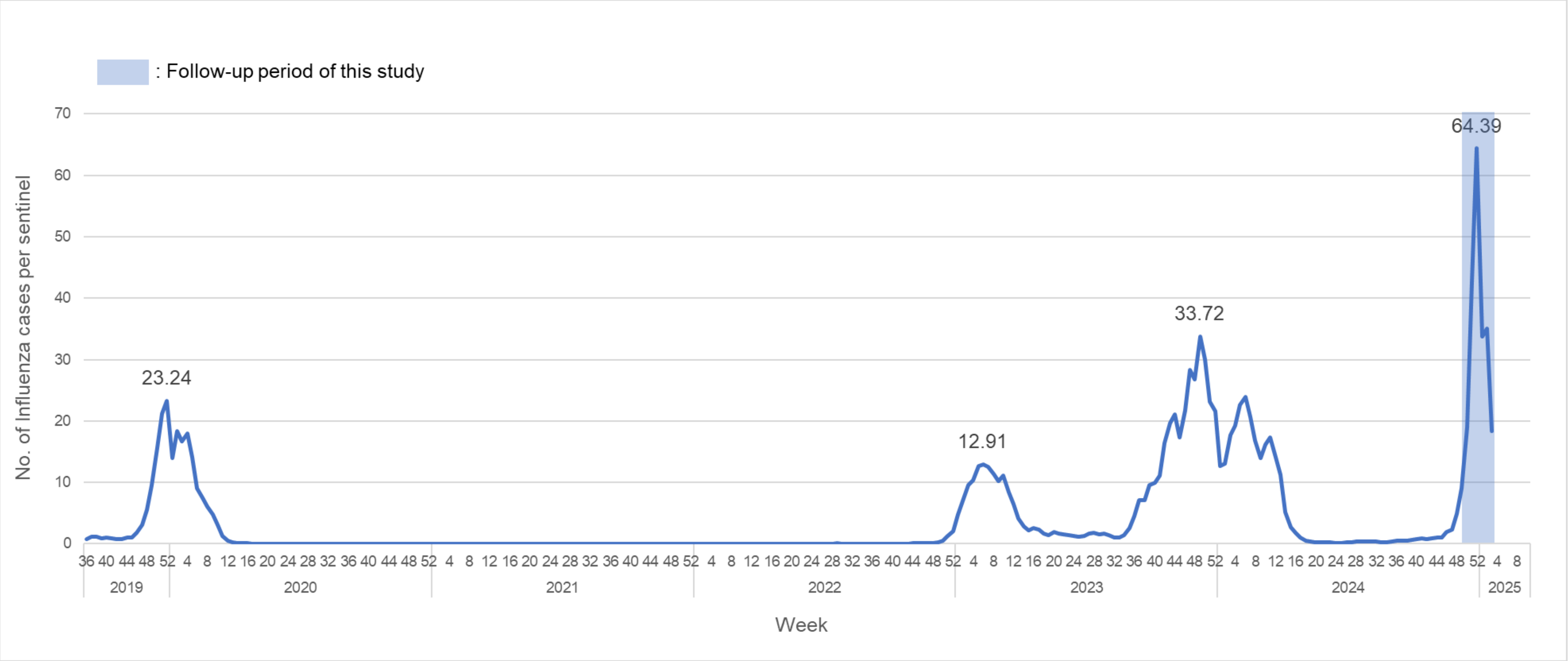


**Supplemental Figure 1**. Number of influenza cases per sentinel in Japan from the 2019/2020 to 2024/2025 seasons.

This figure is based on surveillance data from the Infectious Disease Weekly Report (IDWR), provided by the National Institute of Infectious Diseases, Japan, covering the period from Week 36 of 2019 to Week 3 of 2025. https://www.niid.go.jp/niid/en/data.html

Data labels indicate the peak weekly number of influenza cases per sentinel for each season. The blue bars represent the study follow-up period (December 2, 2024, to January 15, 2025).

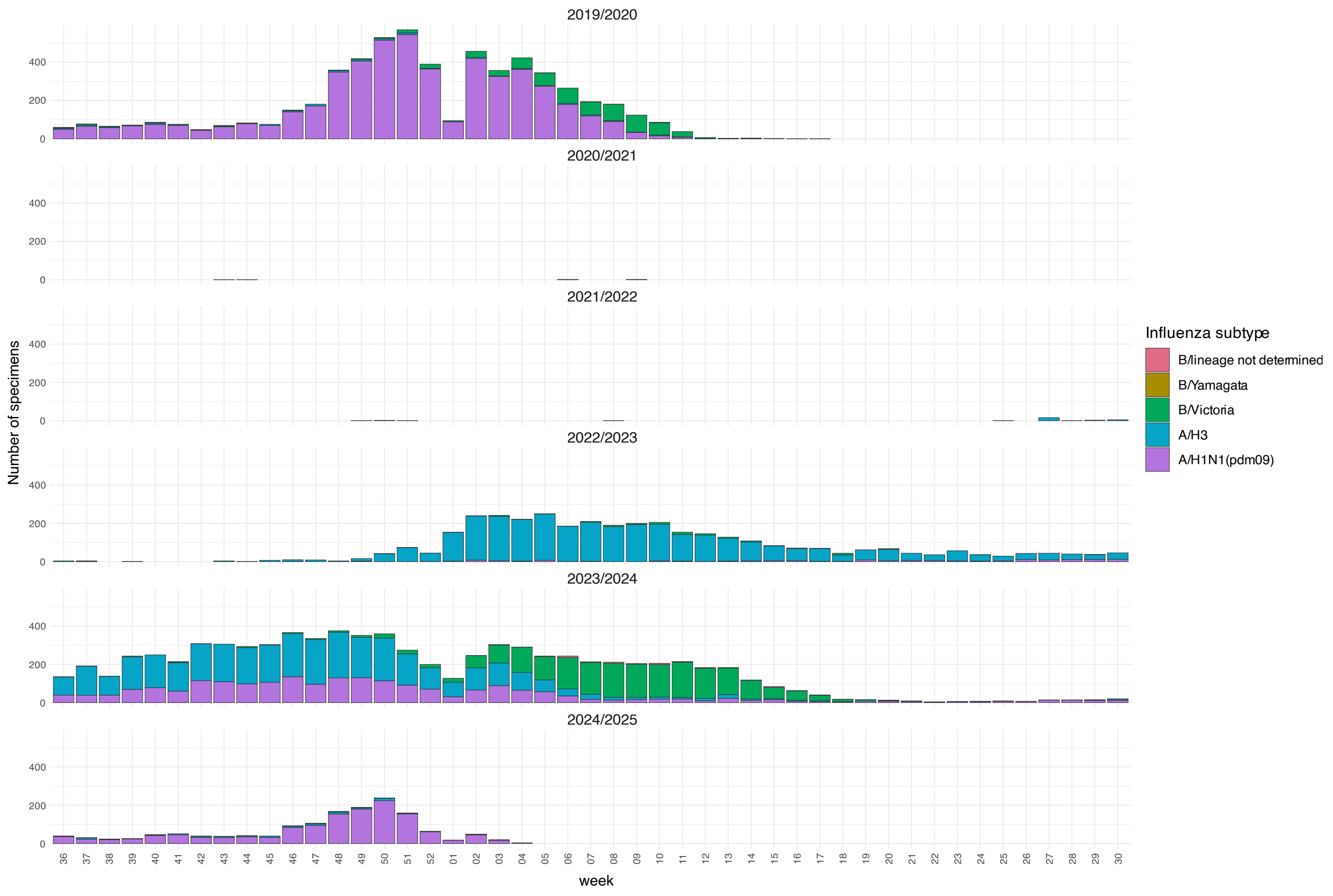


**Supplemental Figure 2**. Number of detected cases based on the influenza subtype in Japan for the 2019/2020 to 2024/2025 seasons.

We collected data on influenza virus isolations and detections from the 2019/2020 to 2024/2025 seasons in Japan from the Infectious Agents Surveillance Report (IASR) published by the NIID (https://www.niid.go.jp/niid/ja/iasr-inf.html) as of January 28, 2025.

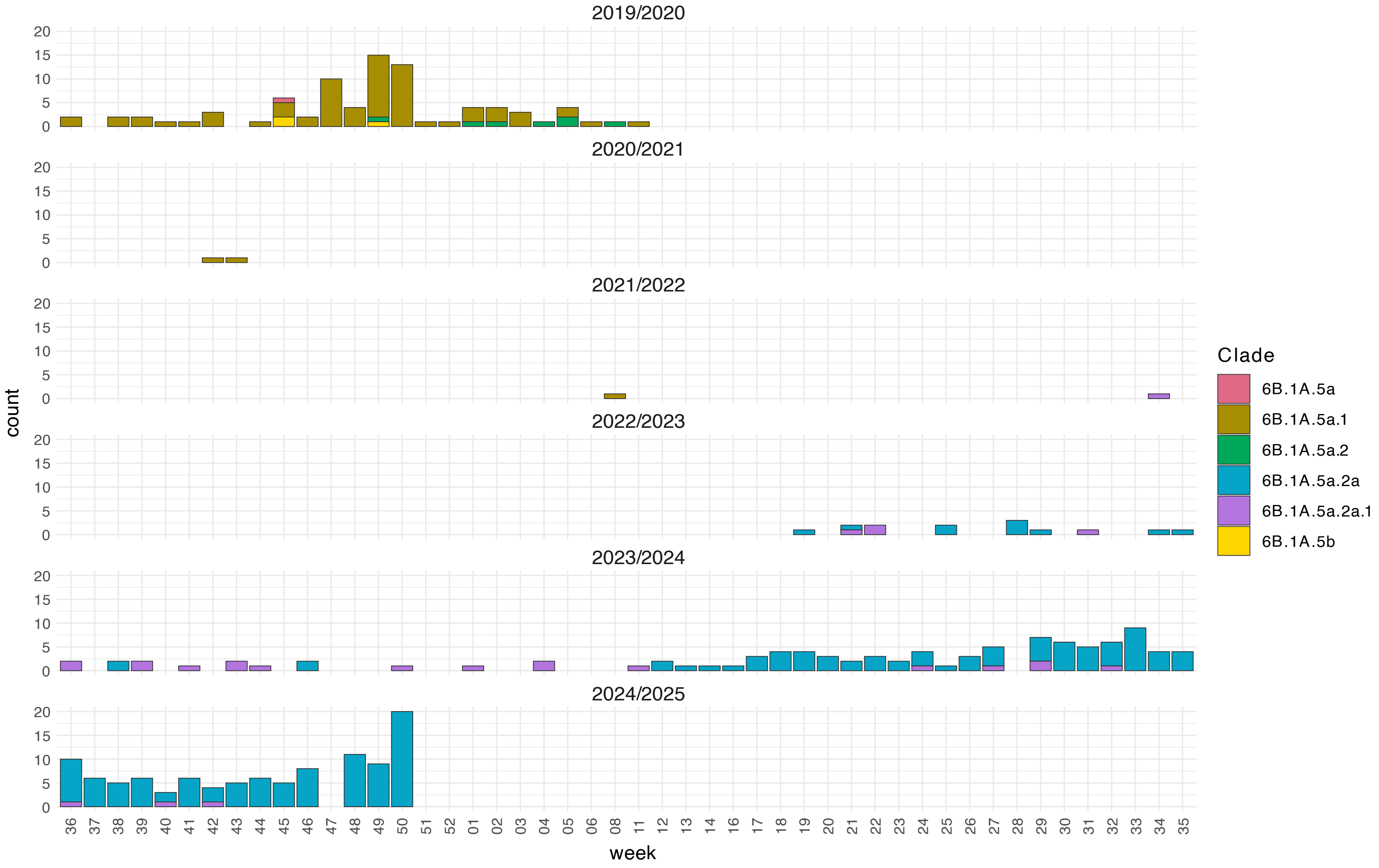


**Supplemental Figure 3**. Number of detected Influenza "A/H1N1" cases based on the clade in Japan for the 2019/2020 to 2024/2025 seasons. The A/H1N1 clade distribution from the 2019/2020 to the 2024/2025 seasons in Japan was analyzed using domestic data from the Global Initiative on Sharing All Influenza Data (GISAID) EpiFlu database (https://gisaid.org/) as of January 27, 2025.

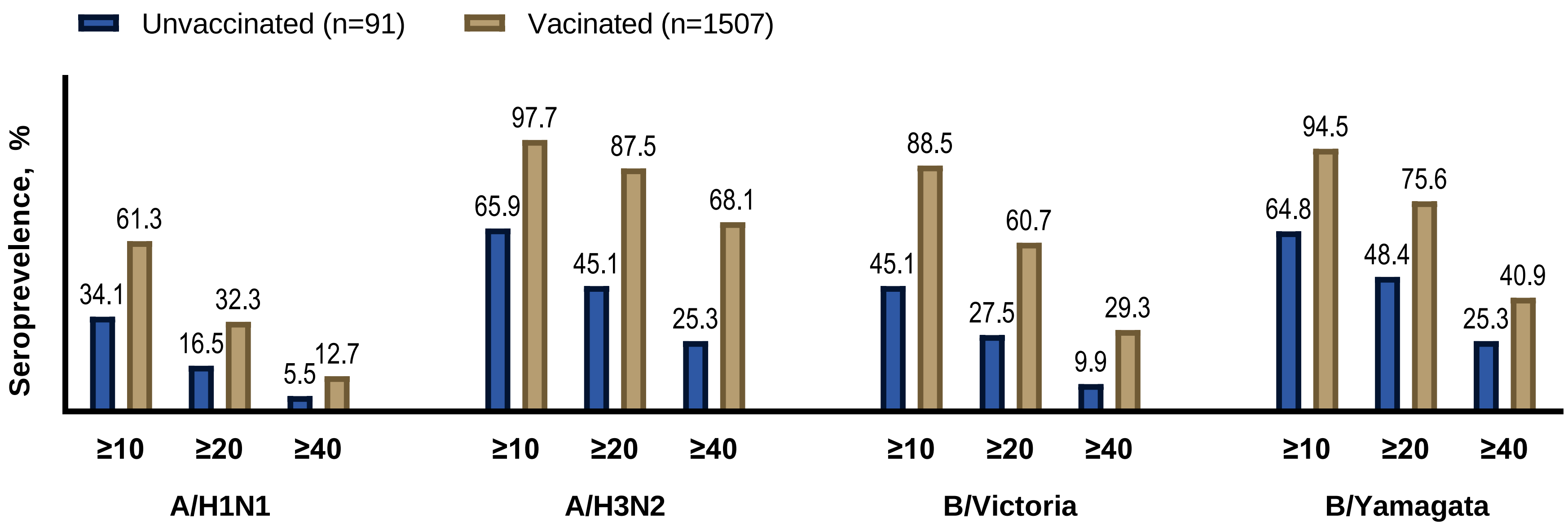


**Supplemental Figure 4**. Seroprevalence of influenza antibodies for HI≥10, HI≥20, and HI≥40, stratified by vaccination status.
Abbreviations: HI, hemagglutination inhibition.
